# Impact of the COVID-19 Pandemic, Stratified by Transfer and COVID-19 Infection Status, on Inpatient Mortality in ST-Elevation Myocardial Infarction Patients, Using a Nationally Representative Database

**DOI:** 10.1101/2023.04.27.23289238

**Authors:** Ahmad Gill, Omar Al-Taweel, Blaine Massey, Salman Mohammed, Jie Ren, Yousif Al-Baghdadi, Akash Parida, Sadaf Fakhra, Osman Rahimi, Tajinder Badial, Saikrishna Patibandla, Tracy Wineinger, Deya Alkhatib, Mohamad Mubder, Chowdhury Ahsan

**Affiliations:** Department of Internal Medicine, Kirk Kerkorian School of Medicine at the University of Nevada, Las Vegas, Las Vegas, Nevada; Department of Cardiology, Kirk Kerkorian School of Medicine at the University of Nevada, Las Vegas, Las Vegas, Nevada; Kirk Kerkorian School of Medicine at the University of Nevada, Las Vegas, Las Vegas, Nevada; Department of Cardiology, Mount Sinai-Brooklyn Hospital, Brooklyn, New York; Kansas College of Osteopathic Medicine, Wichita, Kansas; Department of Cardiology, University of Tennessee Health Science Center, Memphis, Tennessee; Department of Internal Medicine, University Medical Center, Las Vegas, Nevada

**Author notes:** Corresponding Author Name: Ahmad Gill, Affiliation: Department of Internal Medicine, Kirk Kerkorian School of Medicine at the University of Nevada, Las Vegas, Las Vegas, Nevada, Mailing Address: 2040 West Charleston Boulevard Las Vegas, Nevada 89102.

**Keywords:** ST-Elevation Myocardial Infarction, STEMI, COVID-19, Transfer Status

## Abstract

**Introduction:** The coronavirus disease 2019 (COVID-19) pandemic has impacted various aspects of healthcare, including the management of ST-elevation myocardial infarction (STEMI) patients. Our study investigates the in-hospital outcomes and the impact of transfer and COVID-19 infection status on mortality in STEMI patients.

**Methods:** We conducted a retrospective cohort study to compare the inpatient outcomes of STEMI patients in 2020 with STEMI patients from 2016 to 2019 using the National Inpatient Sample database. We performed 1:1 greedy nearest neighbor matching and utilized logistic regression to compare mortality.

**Results:** In our matched cohort, there was no difference in overall mortality between STEMI patients in 2020 and those from 2016 to 2019 (OR 1.00, 95% CI: 0.94-1.05; p = 0.87). When stratified by COVID-19 infection status, regularly admitted STEMI patients with concurrent COVID-19 infection in 2020 had 2.11 times higher odds of inpatient mortality compared to regularly admitted STEMI patients from 2016 to 2019 (OR 2.11, 95% CI: 1.55-2.87; p < 0.001). STEMI acute care transfers with concurrent COVID-19 infection in 2020 had 3.17 times higher odds of inpatient mortality than those from 2016 to 2019 (OR 3.17, 95% CI: 1.83-5.50; p < 0.001). STEMI non-acute care transfers with concurrent COVID-19 infection in 2020 had 5.13 times higher odds of inpatient mortality than those from 2016 to 2019 (OR 5.13, 95% CI: 1.87-14.06; p = 0.001).

**Conclusion:** The COVID-19 pandemic exacerbated many longstanding disparities within our healthcare system. Moving forward, it is crucial to engage in further discussions addressing the national physician shortage, the patient transfer system and healthcare in underserved regions.

## Introduction

The coronavirus disease 2019 (COVID-19) pandemic has impacted various aspects of healthcare, including the management of ST-elevation myocardial infarction (STEMI) patients. Previous studies have demonstrated that patients hospitalized for an acute myocardial infarction with concurrent COVID-19 infection experience higher in-hospital mortality rates compared to those without COVID-19.^1,2^ During the initial pandemic surge, there was a significant decrease in STEMI admissions, which may be attributed to patients avoiding hospitals due to fear of exposure to COVID-19.^3-5^ Moreover, door-to-balloon angioplasty time increased at the onset of the pandemic in numerous countries, including the United States (U.S.), as a result of restructured treatment protocols, stringent infection-control policies and overwhelmed emergency departments.^6-11^ Despite these findings, there is limited research utilizing a nationally representative database to examine the impact of the COVID-19 pandemic on inpatient outcomes of STEMI patients during the first full year of the pandemic. Therefore, our study aims to fill this gap by investigating the in-hospital outcomes and the impact of transfer and COVID-19 infection status on mortality in STEMI patients. We hypothesized that patients with STEMI and concurrent COVID-19 infection in 2020, irrespective of transfer status, will have increased odds of suffering in-hospital mortality compared to STEMI patients without COVID-19 between 2016 to 2019.

## Methods

We conducted a retrospective cohort study to compare the inpatient outcomes of STEMI patients in 2020 with STEMI patients from 2016 to 2019 using the National Inpatient Sample (NIS) database. The NIS is a stratified sample of all-payer inpatient hospital stays in the United States. Each annual dataset contains approximately 7 million hospital stays which, when adjusted for discharge weight, estimates more than 35 million hospitalizations nationally.^12^ The identified discharge record includes one primary diagnosis and up to 29 secondary diagnoses, employing the International Classification of Diseases, Tenth Edition, Clinical Modification (ICD-10-CM) codes. We selected 2016 as the starting point of our study period, as it marks the first full calendar year for ICD-10-CM code usage.^12^

Our study population comprised hospitalized patients aged 18 years or older. Transfer status was determined using the NIS data element “TRAN_IN,” indicating patients admitted as regular hospital admissions or transferred from acute or non-acute care centers. We identified hospitalizations with a primary diagnosis of STEMI and a secondary diagnosis of COVID-19 infection using ICD-10-CM codes listed in the Appendix. Our primary outcome of interest was the in-hospital mortality rate among STEMI patients in 2020 compared to those from 2016 to 2019. Secondary study outcomes included mortality by transfer status, mortality by COVID-19 infection status, percutaneous coronary intervention (PCI) before discharge and overall inpatient complications.

We employed Pearson ***χ***^2^ test to compare categorical variables, while Regular Student’s t-test was used to compare normally distributed continuous variables in our unmatched cohort. Logistic regression was then utilized to calculate a propensity score based on patient demographics (age, race, sex and health insurance), comorbidities (coronary artery disease, type 2 diabetes mellitus, chronic kidney disease, hypertension, obesity, prior myocardial infarction, atrial fibrillation and chronic obstructive pulmonary disease) and hospital factors (region, location and bed size). We evaluated covariate balance using the standardized mean difference (SMD), considering an SMD of less than 0.1 appropriate. We performed 1:1 greedy nearest neighbor matching, setting the caliper at 0.2, to match STEMI patients from 2020 to those from 2016 to 2019 based on propensity scores.

In our matched cohort, we employed multivariable logistic regression to compare mortality. We report the final effect size as an odds ratio (OR) for binary variables. Our two-tailed analysis had a significance threshold set at p < 0.05. We conducted the study using STATA Version 17.

## Results

843,910 patients met our inclusion criteria, with 158,570 (18.8%) patients experiencing a STEMI in 2020 and 685,340 (81.2%) patients experiencing a STEMI between 2016 and 2019. In our unmatched cohort, STEMI patients in 2020 had higher rates of obesity (17.6% vs. 14.5%, p < 0.001) and prior myocardial infarction (10.5% vs. 9.9%, p < 0.001) but decreased rates of atrial fibrillation (9.8% vs. 12.1%, p < 0.001), chronic kidney disease (10.7% vs. 11.2%, p < 0.001), chronic obstructive pulmonary disease (7.6% vs. 8.3%, p < 0.001), hypertension (43.5% vs. 47.2%, p < 0.001), and type 2 diabetes mellitus (15.2% vs. 17.2%, p < 0.001, Table 1).

**Table 1.** Baseline Characteristics of STEMI Patients Before Propensity Score Matching

|  | Year 2020 | Years 2016 to 2019 | p-value |
| --- | --- | --- | --- |
| **n** | 158,570 | 685,340 |  |
| Regular Hospital Admissions | 127,480 (80.4) | 551,195 (80.8) |  |
| Acute Care Transfers | 27,310 (17.2) | 115,330 (16.5) |  |
| Non-Acute Care Transfers | 3780 (2.4) | 18,815 (2.7) |  |
| **Age (mean)** | 63.6 | 63.6 | 0.89 |
| **Sex (%)** |  |  | 0.78 |
| Male | 110,206 (69.5) | 475,626 (69.4) |  |
| Female | 48,364 (30.5) | 209,714 (30.6) |  |
| **Race (%)** |  |  | 0.88 |
| Asian | 4916 (3.1) | 20,560 (3.0) |  |
| Black | 14,271 (9.0) | 60,310 (8.8) |  |
| Hispanic | 13,796 (8.7) | 58,939 (8.6) |  |
| Native American | 793 (0.5) | 4112 (0.6) |  |
| Other | 6026 (3.8) | 24,672 (3.6) |  |
| White | 118,928 (75.0) | 517,432 (75.5) |  |
| **Median Household Income by Quartile (%)** |  |  | 0.24 |
| Quartile 1 (\$1-47,999) | 44,559 (28.1) | 194,637 (28.4) | |
| Quartile 2 (\$48,000-60,999) | 45,351 (28.6) | 187,098 (27.3) | |
| Quartile 3 (\$61,000-81,999) | 37,105 (23.4) | 167,908 (24.5) | |
| Quartile 4 (\$82,000+) | 31,714 (20.0) | 135,697 (19.8) | |
| **APR-DRG Risk Mortality Score<sup>a</sup> (%)** |  |  | <0.001 |
| Score = 0-1 (Minor) | 39,643 (25.0) | 180,930 (26.4) |  |
| Score = 2 (Moderate) | 57,561 (36.3) | 245,352 (35.8) |  |
| Score = 3 (Major) | 26,481 (16.7) | 119,249 (17.4) |  |
| Score = 4 (Extreme) | 34,885 (22.0) | 140,495 (20.5) |  |
| **Payer Status (%)** |  |  | 0.21 |
| Medicare | 70,722 (44.6) | 308,403 (45.0) |  |
| Medicaid | 17,284 (10.9) | 71,961 (10.5) |  |
| Private Insurance | 52,962 (33.4) | 232,330 (33.9) |  |
| Self-Pay | 10,941 (6.9) | 46,603 (6.8) |  |
| No Charge | 951 (0.6) | 4112 (0.6) |  |
| Other | 5709 (3.6) | 22,616 (3.3) |  |
| **Hospital Region (%)** |  |  | 0.99 |
| Northeast | 26,481 (16.7) | 115,822 (16.9) |  |
| Midwest or North Central | 35,678 (22.5) | 155,572 (22.7) |  |
| South | 64,538 (40.7) | 275,507 (40.2) |  |
| West | 31,873 (20.1) | 138,439 (20.2) |  |
| **Hospital Location & Teaching Status (%)** |  |  | <0.001 |
| Rural | 10,466 (6.6) | 44,547 (6.5) |  |
| Urban Non-Teaching | 28,701 (18.1) | 154,202 (22.5) |  |
| Urban Teaching | 119,403 (75.3) | 487,277 (71.1) |  |
| **Hospital Bed Size (%)** |  |  | 0.10 |
| Small | 28,543 (18.0) | 106,913 (15.6) |  |
| Medium | 46,778 (29.5) | 205,602 (30.0) |  |
| Large | 83,249 (52.5) | 372,825 (54.4) |  |
| **Comorbidities (%)** |  |  |  |
| Atrial Fibrillation | 15,605 (9.8) | 82,770 (12.1) | <0.001 |
| Coronary Artery Disease | 121,145 (76.4) | 525,160 (76.6) | 0.05 |
| Chronic Kidney Disease | 17,010 (10.7) | 77,045 (11.2) | <0.001 |
| Chronic Obstructive Pulmonary Disease | 12,085 (7.6) | 57,050 (8.3) | <0.001 |
| Hypertension | 68,980 (43.5) | 323,730 (47.2) | <0.001 |
| Obesity | 27,860 (17.6) | 99,585 (14.5) | <0.001 |
| Previous Myocardial Infarction | 16,715 (10.5) | 67,910 (9.9) | <0.001 |
| Type 2 Diabetes Mellitus | 24,100 (15.2) | 117,790 (17.2) | <0.001 |

<sup>a</sup>APR-DRG scores are calculated from discharge billing codes and based on discharge diagnosis, pre-existing medical conditions and age.

After propensity-score matching, 30,470 patients were included in each cohort. The 2020 STEMI cohort consisted of 24,612 regular hospital admissions, 5151 acute care transfers and 707 non-acute care transfers. The 2016-2019 STEMI cohort consisted of 24,625 regular hospital admissions, 5017 acute care transfers and 828 non-acute care transfers (Table 2).

**Table 2.** Baseline Characteristics of STEMI Patients After Propensity Score Matching

|  | **Year 2020** | **Years 2016 to 2019** | **p-value** |
| --- | --- | --- | --- |
| **n** | 30,470 | 30,470 |  |
| Regular Hospital Admissions | 24,612 (80.8) | 24,625 (80.8) |  |
| Acute Care Transfers | 5151 (16.9) | 5017 (16.5) |  |
| Non-Acute Care Transfers | 707 (2.3) | 828 (2.7) |  |
| **Age (mean)** | 63.6 | 63.8 | 0.20 |
| **Sex (%)** |  |  | 0.04 |
| Male | 21,138 (69.4) | 21,376 (70.2) |  |
| Female | 9332 (30.6) | 9094 (29.8) |  |
| **Race (%)** |  |  | 0.003 |
| Asian | 946 (3.1) | 806 (2.7) |  |
| Black | 2738 (9.0) | 2369 (7.8) |  |
| Hispanic | 2642 (8.7) | 2429 (8.0) |  |
| Native American | 155 (0.5) | 147 (0.5) |  |
| Other | 1148 (3.8) | 1162 (3.8) |  |
| White | 22,841 (75.0) | 23,557 (77.3) |  |
| **Median Household Income by Quartile (%)** |  |  | 0.27 |
| Quartile 1 (\$1-47,999) | 8409 (28.1) | 8198 (27.4) | |
| Quartile 2 (\$48,000-60,999) | 8562 (28.6) | 8280 (27.7) | |
| Quartile 3 (\$61,000-81,999) | 7007 (23.4) | 7377 (24.6) | |
| Quartile 4 (\$82,000+) | 5990 (20.0) | 6095 (20.4) | |
| **APR-DRG Mortality Score<sup>a</sup> (%)** |  |  |  |
| Score = 0-1 (Minor) | 7619 (25.0) | 8049 (26.4) |  |
| Score = 2 (Moderate) | 11,044 (36.3) | 10,803 (35.5) |  |
| Score = 3 (Major) | 5145 (16.9) | 5177 (17.0) |  |
| Score = 4 (Extreme) | 6662 (21.9) | 6441 (21.1) |  |
| **Payer Status (%)** |  |  | <0.001 |
| Medicare | 13,660 (44.8) | 13,918 (45.7) |  |
| Medicaid | 3312 (10.9) | 3148 (10.3) |  |
| Private Insurance | 10,138 (33.3) | 10,402 (34.1) |  |
| Self-Pay | 2090 (6.9) | 1992 (6.5) |  |
| No Charge | 187 (0.6) | 149 (0.5) |  |
| Other | 1083 (3.6) | 861 (2.8) |  |
| **Hospital Region (%)** |  |  | 0.99 |
| Northeast | 5146 (16.9) | 5176 (17.0) |  |
| Midwest or North Central | 6713 (22.0) | 6698 (22.0) |  |
| South | 12,557 (41.2) | 12,516 (41.0) |  |
| West | 6054 (19.9) | 6080 (20.0) |  |
| **Hospital Location & Teaching Status (%)** |  |  | 0.14 |
| Rural | 1976 (6.5) | 1661 (5.4) |  |
| Urban Non-Teaching | 5528 (18.1) | 6026 (19.8) |  |
| Urban Teaching | 22,966 (75.4) | 22,783 (74.8) |  |
| **Hospital Bed Size (%)** |  |  | 0.62 |
| Small | 5454 (17.9) | 5129 (16.8) |  |
| Medium | 9028 (29.6) | 9312 (30.6) |  |
| Large | 15,988 (52.5) | 16,029 (52.6) |  |
| **Comorbidities (%)** |  |  |  |
| Atrial Fibrillation | 3015 (9.9) | 2987 (9.8) | 0.70 |
| Coronary Artery Disease | 23,279 (76.4) | 23,405 (76.8) | 0.27 |
| Chronic Kidney Disease | 3305 (10.8) | 3054 (10.0) | <0.001 |
| Chronic Obstructive Pulmonary Disease | 2328 (7.6) | 2129 (7.0) | 0.002 |
| Hypertension | 13,248 (43.5) | 13,450 (44.1) | 0.10 |
| Obesity | 5370 (17.6) | 5265 (17.3) | 0.26 |
| Previous Myocardial Infarction | 3233 (10.6) | 3055 (10.0) | 0.02 |
| Type 2 Diabetes Mellitus | 4623 (15.2) | 4404 (14.5) | 0.01 |

<sup>a</sup>APR-DRG scores are calculated from discharge billing codes and based on discharge diagnosis, pre-existing medical conditions and age.

In our matched cohorts, there was no difference in overall mortality between STEMI patients in 2020 compared to STEMI patients from 2016 to 2019 (OR 1.00, 95% CI: 0.94-1.05; p = 0.87). However, STEMI patients in 2020 had 1.06 times higher odds of PCI than those in 2016-2019 (OR 1.06, 95% CI: 1.03-1.08; p < 0.001). There was no significant difference in mean time to PCI between our two cohorts (0.1 days vs. 0.1 days, p = 0.44). When stratified by COVID-19 infection status, STEMI patients with concurrent COVID-19 infection in 2020 had 2.44 times higher mortality odds than STEMI patients without COVID-19 from 2016 to 2019 (OR 2.44, 95% CI: 1.89-3.15; p < 0.001).

In our matched cohorts, when separated by transfer status, no significant difference in overall mortality was observed between regularly admitted, non-transferred STEMI patients in 2020 and those from 2016 to 2019 (OR 0.97, 95% CI: 0.91-1.04; p = 0.43). Regularly admitted STEMI patients in 2020 had 1.06 times higher odds of undergoing PCI than those in 2016-2019 (OR 1.06, 95% CI: 1.03-1.09; p < 0.001). There was no significant difference in mean time to PCI between our two regular admission cohorts (0.1 days vs. 0.1 days, p = 0.64). When stratified by COVID-19 infection status, regularly admitted STEMI patients with concurrent COVID-19 infection in 2020 had 2.11 times higher odds of mortality compared to regularly admitted STEMI patients from 2016 to 2019 (OR 2.11, 95% CI: 1.55-2.87; p < 0.001, Figure 1).

**Figure 1:**
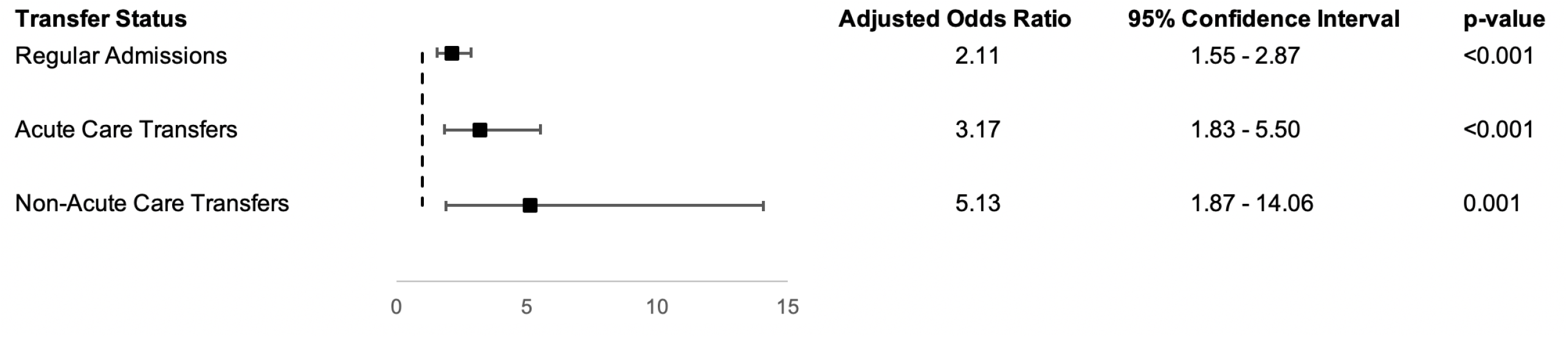
The Effect of Concurrent COVID-19 Infection on Mortality by Transfer Status in STEMI Patients After Propensity Score Matching

No significant difference in overall mortality was found for STEMI patients transferred from acute care centers in 2020 compared to those admitted as acute care transfers from 2016 to 2019 (OR 1.12, 95% CI: 0.97-1.28; p = 0.11). Similarly, there was no significant difference in odds of PCI (OR 1.01, 95% CI: 0.95-1.07; p = 0.73) and mean time to PCI between our two acute care transfer cohorts (0.2 days vs. 0.3 days, p = 0.46). When stratified by COVID-19 infection status, STEMI acute care transfers with concurrent COVID-19 infection in 2020 had 3.17 times higher odds of inpatient mortality than those from 2016 to 2019 (OR 3.17, 95% CI: 1.83-5.50; p < 0.001, Figure 1).

No significant difference in overall inpatient mortality was found for STEMI patients transferred from non-acute care centers in 2020 compared to STEMI non-acute care transfers from 2016 to 2019 (OR 0.94, 95% CI: 0.69-1.27; p = 0.68). Non-acute care STEMI transfers in 2020 had 1.26 times higher odds of PCI than those from 2016 to 2019 (OR 1.26, 95% CI: 1.07-1.47; p = 0.005). There was no significant difference in mean time to PCI between our two non-acute care transfer cohorts (0.1 days vs. 0.1 days, p = 0.96). When stratified by COVID-19 infection status, STEMI non-acute care transfers with concurrent COVID-19 infection in 2020 had 5.13 times higher odds of inpatient mortality than those from 2016 to 2019 (OR 5.13, 95% CI: 1.87-14.06; p = 0.001, Figure 1).

Compared to STEMI patients from 2016 to 2019, STEMI patients in 2020 were more likely to have acute hepatic failure (OR 1.23, 95% CI: 1.15-1.32; p < 0.001), acute kidney injury (OR 1.19, 95% CI: 1.15-1.23; p < 0.001), acute respiratory distress syndrome (OR 1.52, 95% CI: 1.17-1.97; p = 0.001), atrial flutter (OR 1.90, 95% CI: 1.35-2.68; p < 0.001), hypothyroidism (OR 1.42, 95% CI: 1.17-1.73; p < 0.001), viral cardiomyopathy (OR 4.32, 95% CI: 1.08-17.28; p = 0.04), severe sepsis (OR 1.28, 95% CI: 1.16-1.42; p < 0.001), supraventricular tachycardia (OR 1.13, 95% CI: 1.04-1.23; p = 0.003) and ventricular tachycardia (OR 1.22, 95% CI: 1.18-1.26; p < 0.001). However, they were less likely to have postprocedural acute kidney injury (OR 0.31, 95% CI: 0.10-0.99; p = 0.04) and postprocedural respiratory failure (OR 0.62, 95% CI: 0.50-0.77; p < 0.001) during their inpatient admission (Table 3).

**Table 3.** Inpatient Complications Comparing STEMI Patients in 2020 to those between 2016 and 2019 after Propensity Score Matching

| **Outcomes** | **OR (95% CI)** | **p-value** |
| --- | --- | --- |
| Atrial Fibrillation | 0.96 (0.91-1.01) | 0.13 |
| Atrial Flutter | 1.90 (1.35-2.68) | <0.001 |
| Acute Hepatic Failure | 1.23 (1.15-1.32) | <0.001 |
| Acute Kidney Injury | 1.19 (1.15-1.23) | <0.001 |
| Acute Respiratory Distress Syndrome | 1.52 (1.17-1.97) | 0.001 |
| Cardiogenic Shock | 1.12 (0.88-1.44) | 0.36 |
| Hypertension | 1.85 (0.48-7.16) | 0.37 |
| Hypotension | 1.04 (0.88-1.24) | 0.63 |
| Hypothyroidism | 1.42 (1.17-1.73) | <0.001 |
| Fever | 1.38 (0.84-2.25) | 0.20 |
| Myocarditis | 1.53 (0.79-2.95) | 0.21 |
| Pneumothorax | 1.01 (0.70-1.44) | 0.98 |
| Ventricular Fibrillation | 1.01 (0.97-1.06) | 0.58 |
| Viral Cardiomyopathy | 4.32 (1.08-17.28) | 0.04 |
| Severe Sepsis | 1.28 (1.16-1.42) | <0.001 |
| Supraventricular Tachycardia | 1.13 (1.04-1.23) | 0.003 |
| Ventricular Tachycardia | 1.22 (1.18-1.26) | <0.001 |

## Discussion

Our study focused on comparing the impact of the COVID-19 pandemic on transfer status and mortality in STEMI patients. After propensity score matching and stratification by transfer status, no difference in mortality was found in STEMI patients in 2020 compared to those from 2016 to 2019. However, STEMI patients with concurrent COVID-19 infection in 2020 had higher odds of mortality across all three cohorts (regular admissions, acute care transfers and non-acute care transfers) than STEMI patients without COVID-19 from 2016 to 2019. STEMI patients in 2020 were also more likely to have acute hepatic failure, acute respiratory distress syndrome, atrial flutter, hypothyroidism, viral cardiomyopathy, severe sepsis, supraventricular tachycardia and ventricular tachycardia.

To our knowledge, our study is the largest study examining the impact of the COVID-19 pandemic on STEMI mortality while using STEMI patients between 2016 to 2019 as a baseline for comparison. A significant decrease in STEMI admissions was registered globally during the initial stages of the COVID-19 pandemic. Reasons for the declining incidence of STEMI include stay-at-home orders, a decrease in activities that could trigger STEMI and fear of contracting COVID-19.^13-15^ Additionally, several studies in European countries observed that the pandemic was associated with a significantly longer ischemia time, including a longer door-to-balloon time and a significant reduction in the number of STEMI patient undergoing PCI.^16,17^ Factors associated with delayed management include an increase in transportation time, the need for personal protection equipment and the increased burden on healthcare systems overall.^17^ As 2020 progressed, overall STEMI mortality decreased, which is why there is no significant difference in overall STEMI mortality in 2020 compared to STEMI mortality between 2016 to 2019.^1^

Consistent with our findings, a study by Goel et al. also found that concurrent COVID-19 infection increases the odds of inpatient STEMI mortality.^1^ Previous studies have demonstrated that COVID-19 infection raises the risk of acute myocardial infarction through various mechanisms, including a cytokine storm, hypercoagulability and endothelial dysfunction.^18-25^ Although the specific factors contributing to inferior inpatient outcomes for STEMI patients with concurrent COVID-19 infection are not yet fully understood, it is speculated that these patients are more susceptible to associated cardiovascular complications and carry a higher thrombus burden.^1^

Our study is also the first to examine the impact of concurrent COVID-19 infection on STEMI mortality stratified by transfer status. Regardless of transfer status, STEMI patients with concurrent COVID-19 faced higher mortality odds than those without COVID-19. Although hospitals often transfer only the sickest patients, the pronounced difference in mortality odds between acute and non-acute care transfer cohorts relative to the regular hospital admission cohort suggests that the lack of healthcare access, particularly in underserved regions, may also be a contributing factor. Approximately 60 million individuals reside in rural America. On average, they are older, have higher rates of chronic medical conditions than urban residents and rely on local hospitals for healthcare.^26-28^ Unfortunately, since 2005, 161 rural hospitals have closed. As of February 2019, 673 rural hospitals were at risk of closing due to lower patient volumes, workforce shortages, inadequate reimbursements and costly medications.^29-31^ Moreover, as of 2018, the United States had only 19.1 total hospitals per 1,000,000 population, significantly lower than the comparable country average of 32.7.^32^ The COVID-19 pandemic further exacerbated these issues, with more than 36 hospitals entering bankruptcy and 21 hospitals closing in 2020.^33,34^ Additionally, nursing homes confronted with severe COVID-19 outbreaks in 2020 experienced significant reductions in nursing staff levels and struggled to recover the lost staff even after 16 weeks.^35^ According to the Bureau of Labor Statistics, nursing homes have lost nearly 229,000 caregivers (14% of their workforce) since February 2020. These factors potentially contributed to delays in patient care, resulting in increased complications and mortality.^36^

Another compounding factor for increased STEMI and COVID-19 mortality is the national physician shortage. The U.S. Census Bureau estimated an increase of 13,787,044 individuals in the 65-and-older population between 2010 and 2020.^37^ In 2012, the Association of American Medical Colleges (AAMC) predicted that by 2020, the United States would have a shortage of 45,400 primary care physician and 46,100 medical specialists.^38,39^ Unfortunately, the COVID-19 pandemic has exacerbated this issue by causing increased healthcare worker burnout, trauma and mental health issues.^40,41^ Between the beginning of 2019 and the fall of 2021, 3272 physicians left the workforce. Since the start of the pandemic, 3500 healthcare workers have died, with 17% being physicians.^40-42^ The American population is projected to grow from 328 million in 2019 to 363 million in 2034, with a 42.4% increase in individuals aged 65 and above.^38^ During this period, the United States could experience an estimated physician shortage ranging from 37,800 to 124,000, further intensifying disparities in healthcare access.^38,42^

STEMI patients with concurrent COVID-19 infection constitute a high-risk cohort with higher odds of facing inpatient mortality than STEMI patients without COVID-19. The national physician shortage and unequal distribution of healthcare resources may place many patients who lack appropriate healthcare access at increased risk for inpatient mortality. Further discussion and research should explore potential interventions to address these challenges, thereby better equipping our healthcare system to manage not only present issues but also future global health challenges.

Our study has limitations. The use of the NIS database introduces an inherent risk of miscoded diagnoses. Clinically relevant data, such as left ventricular ejection fraction, imaging results, medications administered, route of PCI access, number of stents placed, procedural details, outcomes after discharge, readmission rate and prior hospitalizations, were unavailable, potentially causing confounding and impacting the results. Of the 158,570 STEMI patients in 2020, 30,470 (19.2%) patients were included in the final analysis. Patient selection may change based on calculated propensity scores, introducing potential selection bias and affecting the generalizability of our findings. Lastly, the type of COVID-19 variant and the relationship between vaccination status and in-hospital outcomes could not be assessed.

## Conclusion

In our nationally representative sample of inpatient hospitalizations, STEMI patients in 2020 did not exhibit higher odds of inpatient mortality, even when separated by transfer status. However, when stratified by COVID-19 infection status, STEMI patients with concurrent COVID-19 infection in 2020 had higher odds of in-hospital mortality than STEMI admissions without COVID-19 from 2016 to 2019. The COVID-19 pandemic exacerbated many longstanding disparities within our healthcare system. Moving forward, it is crucial to engage in further discussions addressing the national physician shortage, the patient transfer system and healthcare in underserved regions.

## Data Availability

The data is taken from the National Inpatient Sample database.

## Funding

This research received no grants from any funding agency in the public, commercial or not-for-profit sectors.

## Disclosure

The authors have no relevant financial or non-financial interests to disclose.

### Appendix

<u>STEMI ICD-10 Codes</u>

I21.01, I21.02, I21.09, I21.11, I21.19, I21.21, I21.29, I21.3

<u>COVID-19 ICD-10 Code</u>

U07.1

## Notes

### Competing Interest Statement

The authors have declared no competing interest.

### Funding Statement

No authors received any payment or services from a third party.

## References

1. Goel A, Malik AH, Bandyopadhyay D, et al. Impact of COVID-19 on outcomes of patients hospitalized with STEMI: A nationwide propensity-matched analysis. Current Problems in Cardiology. 2023;48(4):101547. doi:10.1016/j.cpcardiol.2022.101547

2. Baral N, Abusnina W, Balmuri S, et al. COVID-19 Positive Status is Associated with Increased In-hospital Mortality in Patients with Acute Myocardial Infarction: A Systematic Review and Meta-analysis. J Community Hosp Intern Med Perspect. 2022;12(4):17–24. Published 2022 Sep 9. doi:10.55729/2000-9666.1103

3. Lertsanguansinchai P, Chokesuwattanaskul R, Limjaroen T, et al. Clinical characteristics and in-hospital mortality in patients with STEMI during the COVID-19 outbreak in Thailand. Biomedicines. 2022;10(11):2671. doi:10.3390/biomedicines10112671

4. De Rosa S, Spaccarotella C, Basso C, et al. Reduction of hospitalizations for myocardial infarction in Italy in the COVID-19 ERA. European Heart Journal. 2020;41(22):2083–2088. doi:10.1093/eurheartj/ehaa409

5. Saad M, Kennedy KF, Imran H, et al. Association Between COVID-19 Diagnosis and In-Hospital Mortality in Patients Hospitalized With ST-Segment Elevation Myocardial Infarction. JAMA. 2021;326(19):1940–1952. doi:10.1001/jama.2021.18890

6. Chew NWS, Sia C-H, Wee H-L, et al. Impact of the COVID-19 pandemic on door-to-balloon time for primary percutaneous coronary intervention - results from the Singapore Western Stemi Network -. Circulation Journal. 2021;85(2):139–149. doi:10.1253/circj.cj-20-0800

7. Bangalore S, Halista M. STEMI outcomes in the era of COVID-19: Reaffirmation of an unfortunate reality. EuroIntervention. 2021;16(17):1379–1380. doi:10.4244/eijv16i17a250

8. Hammad TA, Parikh M, Tashtish N, et al. Impact of covid -19 pandemic on st-elevation myocardial infarction in a non-covid -19 epicenter. Catheterization and Cardiovascular Interventions. 2020;97(2):208–214. doi:10.1002/ccd.28997

9. Garcia S, Dehghani P, Stanberry L, et al. Trends in clinical presentation, management, and outcomes of STEMI in patients with covid-19. Journal of the American College of Cardiology. 2022;79(22):2236–2244. doi:10.1016/j.jacc.2022.03.345

10. Modin D, Claggett B, Sindet-Pedersen C, et al. Acute covid-19 and the incidence of ischemic stroke and acute myocardial infarction. Circulation. 2020;142(21):2080–2082. doi:10.1161/circulationaha.120.050809

11. Dogan Z, Erden I, Bektasoglu G, Karabulut A. Association between history of polymerase chain reaction-verified COVID-19 infection and outcomes of subsequent st-elevation myocardial infarction. Angiology. 2022:000331972211399. doi:10.1177/00033197221139918

12. Overview of the National (Nationwide) Inpatient Sample (NIS). HCUP. https://www.hcup-us.ahrq.gov/nisoverview.jsp. Accessed July 16, 2022.

13. Solomon MD, McNulty EJ, Rana JS, et al. The Covid-19 Pandemic and the Incidence of Acute Myocardial Infarction. N Engl J Med. 2020;383(7):691–693. doi:10.1056/NEJMc2015630

14. Zitelny E, Newman N, Zhao D. STEMI during the COVID-19 Pandemic - An Evaluation of Incidence. Cardiovasc Pathol. 2020 Sep-Oct;48:107232. doi: 10.1016/j.carpath.2020.107232. Epub 2020 May 1

15. Furnica C, Chistol RO, Chiran DA, et al. The Impact of the Early COVID-19 Pandemic on ST-Segment Elevation Myocardial Infarction Presentation and Outcomes-A Systematic Review and Meta-Analysis. Diagnostics (Basel). 2022;12(3):588. Published 2022 Feb 25. doi:10.3390/diagnostics12030588

16. De Luca G, Verdoia M, Cercek M, et al. Impact of COVID-19 Pandemic on Mechanical Reperfusion for Patients With STEMI. J Am Coll Cardiol. 2020;76(20):2321–2330. doi:10.1016/j.jacc.2020.09.546

17. Roffi M, Guagliumi G, Ibanez B. The Obstacle Course of Reperfusion for ST-Segment-Elevation Myocardial Infarction in the COVID-19 Pandemic. Circulation. 2020;141(24):1951–1953. doi:10.1161/CIRCULATIONAHA.120.047523

18. Esposito L, Cancro FP, Silverio A, et al. Covid-19 and acute coronary syndromes: From pathophysiology to clinical perspectives. Oxidative Medicine and Cellular Longevity. 2021;2021:1–13. doi:10.1155/2021/4936571

19. Masi P, Hékimian G, Lejeune M, et al. Systemic inflammatory response syndrome is a major contributor to covid-19–associated coagulopathy. Circulation. 2020;142(6):611–614. doi:10.1161/circulationaha.120.048925

20. Panigada M, Bottino N, Tagliabue P, et al. Hypercoagulability of Covid-19 patients in Intensive Care Unit: A report of thromboelastography findings and other parameters of hemostasis. Journal of Thrombosis and Haemostasis. 2020;18(7):1738–1742. doi:10.1111/jth.14850

21. Choudry FA, Hamshere SM, Rathod KS, et al. High thrombus burden in patients with covid-19 presenting with st-segment elevation myocardial infarction. Journal of the American College of Cardiology. 2020;76(10):1168–1176. doi:10.1016/j.jacc.2020.07.022

22. Warner SJ, Auger KR, Libby P. Human interleukin 1 induces interleukin 1 gene expression in human vascular smooth muscle cells. Journal of Experimental Medicine. 1987;165(5):1316–1331. doi:10.1084/jem.165.5.1316

23. Warner SJ, Libby P. Human vascular smooth muscle cells. target for and source of tumor necrosis factor. The Journal of Immunology. 1989;142(1):100–109. doi:10.4049/jimmunol.142.1.100

24. Pons S, Arnaud M, Loiselle M, Arrii E, Azoulay E, Zafrani L. Immune consequences of endothelial cells’ activation and dysfunction during sepsis. Critical Care Clinics. 2020;36(2):401–413. doi:10.1016/j.ccc.2019.12.001

25. Xuan Y, Gào X, Holleczek B, Brenner H, Schöttker B. Prediction of myocardial infarction, stroke and cardiovascular mortality with urinary biomarkers of oxidative stress: Results from a large cohort study. International Journal of Cardiology. 2018;273:223–229. doi:10.1016/j.ijcard.2018.08.002

26. Kuehn BM. Hypertension rates in rural areas outpace those in urban locales. JAMA. 2020;323(24):2454. doi:10.1001/jama.2020.9382

27. Adults living in rural counties more likely to be obese than adults living in urban counties. Centers for Disease Control and Prevention. https://www.cdc.gov/nccdphp/dnpao/division-information/media-tools/mmwr-obesity-rural-counties.html#:~:text=Obesity%20prevalence%20was%20significantly%20higher,the%20South%20and%20Northeast%20regions. Published November 12, 2020. Accessed January 14, 2023.

28. Census.gov. https://www.census.gov/content/dam/Census/library/publications/2019/acs/acs-41.pdf. Accessed January 15, 2023.

29. Rural Hospital closures. Sheps Center. https://www.shepscenter.unc.edu/programs-projects/rural-health/rural-hospital-closures/. Published January 11, 2022. Accessed January 14, 2023.

30. Topchik M. Rural Relevance 2017: Assessing the State of Rural Healthcare in America - The Chartis Group. https://www.chartis.com/forum/wp-content/uploads/2017/05/The-Rural-Relevance-Study_2017.pdf. Accessed January 14, 2023.

31. Neel J, Neighmond P. Poll: Many rural Americans struggle with financial insecurity, access to health care. NPR. https://www.npr.org/sections/health-shots/2019/05/21/725059882/poll-many-rural-americans-struggle-with-financial-insecurity-access-to-health-ca. Published May 21, 2019. Accessed January 14, 2023.

32. Hospital resources. Peterson-KFF Health System Tracker. https://www.healthsystemtracker.org/indicator/quality/hospital-resources/#Total%20hospitals%20per%201,000,000%20population,%202018%20or%20nearest%20year. Published November 2, 2022. Accessed January 14, 2023.

33. Fact sheet: Covid-19 pandemic results in bankruptcies or closures for some hospitals: AHA. American Hospital Association. https://www.aha.org/fact-sheets/2020-11-09-fact-sheet-covid-19-pandemic-results-bankruptcies-or-closures-some-hospitals. Accessed January 14, 2023.

34. Ellison A. 21 hospital closures in 2020. Becker’s Hospital Review. https://www.beckershospitalreview.com/finance/21-hospital-closures-in-2020.html. Accessed January 14, 2023.

35. Shen K, McGarry BE, Grabowski DC, Gruber J, Gandhi AD. Staffing patterns in US nursing homes during COVID-19 outbreaks. JAMA Health Forum. 2022;3(7). doi:10.1001/jamahealthforum.2022.2151

36. Historic staffing shortages continue to force nursing homes to limit new admissions, creating bottlenecks at hospitals and reducing access to care for seniors. Press Releases. https://www.ahcancal.org/News-and-Communications/Press-Releases/Pages/Historic-Staffing-Shortages-Continue-To-Force-Nursing-Homes-To-Limit-New-Admissions,-Creating-Bottlenecks-at-Hospitals-and-.aspx. Accessed January 13, 2023.

37. Bureau USC. 65 and older population grows rapidly as baby boomers age. Census.gov. https://www.census.gov/newsroom/press-releases/2020/65-older-population-grows.html. Published October 8, 2021. Accessed January 14, 2023

38. AAMC report reinforces mounting physician shortage. AAMC. http://www.aamc.org/news-insights/press-releases/aamc-report-reinforces-mounting-physician-shortage. Published June 11, 2021. Accessed January 14, 2023.

39. Kirch DG, Henderson MK, Dill MJ. Physician workforce projections in an era of health care reform. Annual Review of Medicine. 2012;63(1):435–445. doi:10.1146/annurev-med-050310-134634

40. HP -2022 13 impact of the COVID-19 pandemic on the hospital and … - aspe. https://aspe.hhs.gov/sites/default/files/documents/9cc72124abd9ea25d58a22c7692dccb6/aspe-covid-workforce-report.pdf. Accessed January 15, 2023.

41. What we’ve learned about covid-19, Burnout and the doctor shortage. American Medical Association. https://www.ama-assn.org/practice-management/sustainability/what-we-ve-learned-about-covid-19-burnout-and-doctor-shortage. Published May 4, 2022. Accessed January 15, 2023.

42. Dill M. We already needed more doctors. then COVID-19 hit. AAMC. https://www.aamc.org/news-insights/we-already-needed-more-doctors-then-covid-19-hit#:~:text=According%20to%20data%20from%20Kaiser,tremendous%20loss%20for%20our%20nation. Published June 17, 2021. Accessed February 20, 2023.

